# Vaccination efforts in Brazil: scenarios and perspectives under a mathematical modeling approach

**DOI:** 10.1101/2021.02.22.21252208

**Authors:** Thomas Nogueira Vilches, Felipe Alves Rubio, Rafael Augusto Forti Perroni, Gabriel Berg de Almeida, Carlos Magno Castelo Branco Fortaleza, Cláudia Pio Ferreira

**Affiliations:** Institute of Mathematics, Statistics and Scientific Computing, UNICAMP, Brazil; Medical School, UNESP, Botucatu Campus, Brazil; Institute of Biosciences, UNESP, Botucatu Campus, Brazil

**Keywords:** agent-based model, behavioral change, immunization scheme

## Abstract

An agent-based model is proposed to access the impact of vaccination strategies to halt the COVID-19 spread. The model is parameterized using data from São Paulo State, Brazil. It was considered the two vaccines that are already approved for emergency use in Brazil, the CoronaVac vaccine developed by the Chinese bio-pharmaceutical company Sinovac and the Oxford-AstraZeneca vaccine (ChadOx1) developed by Oxford University and the British laboratory AstraZeneca. Both of them are two-dose schemes, but the efficacy and the interval between doses are different. We found that even in the worst scenario, in which the vaccine does not prevent infection either severe symptoms, the number of deaths decreases from 122 to 99 for CoronaVac application and to 80 for ChadOx1 administration. The same patterns have been seen in hospitalizations. Nevertheless, we show that when a low risk perception occurs, the reduction values decrease between 2% to 4%. Moreover, the increase of disease prevalence also jeopardizes immunization, showing the importance of the mitigation measures maintenance. On the other hand, doubling the vaccination rate would be able to significantly decrease the disease outcomes, reducing deaths by up to 74.4%. In conclusion, vaccination, along with non-pharmaceutical measures, is key to the control of COVID-19 in Brazil.

## 1 Introduction

Starting between November and December of 2020, the Coronavirus Disease 2019 (COVID-19), caused by the virus SARS-CoV-2, pandemic claimed several lives worldwide. More than fourteen months later, many questions are still not answered, including its origin, routes of transmission, and effective treatment.

After struggling for months to halt or diminish transmission solely through non-pharmacological measures such as quarantine, self-isolation, lockdown, mobility restriction on several spatial scales, a local and global immunization program through vaccination appears as a real possibility. Several vaccines have been approved for emergency use in several countries. They differ by technological and conceptual platforms, grouped based on the protective immune response they trigger [1]. The main platforms are RNA, DNA, Vector (non-replicating), Vector (replicating), Inactivated, Live-attenuated, Protein subunit, and Virus-like particle. Currently, there are about fifty official vaccine projects that reached human experimentation. Ten vaccine platforms are already in use, of which two of them are RNA, three are vector (non-replicating), four are inactivated, and one is subunit [2].

In Brazil, it was not different. On January 17, 2021, the National Health Surveillance Agency (Anvisa) approved the emergency use of the Coronavac vaccine developed by the Chinese biopharmaceutical company Sinovac and the ChadOx1 vaccine developed by Oxford University and the British laboratory AstraZeneca. Both of them performed phase 3 of their study in Brazil within a collaboration with two important Brazilian Institutions: Butantan and the Oswaldo Cruz Foundation (Fiocruz). The efficacy against symptoms presented by Coronavac and Oxford-AstraZeneca vaccines, after two doses, was 50.38% and 70.42%, respectively [3, 4]. However, according to Anvisa, the data presented regarding efficacy had no statistical significance for severe cases, although some protection is expected.

Brazil is known as one of the countries that produce and export vaccines to the rest of the world. The leading factories of Butantan and Manguinhos (Fiocruz) can produce from 1 to 3 million doses per day [5]. Moreover, the country counts with the National Immunization Program (PNI) through the Unified Health System (SUS). The PNI is responsible for successfully eradicating and controlling several endemic diseases in the country, increasing the average Brazilian life expectancy for almost half a century. The robust and complex infrastructure of SUS guarantees access to medicines, vaccines, and healthcare to the population. It provides high-cost medicines, expensive medical procedures, and vaccines to the entire population, including those with private health insurance and people in difficult-to-access areas, such as indigenous people and *quilombolas*^1^ communities [6, 7]. This well-established unified system is an important tool against the COVID-19 epidemics and can easily vaccinate almost one million individuals per day as shown in other immunization campaigned [8]. However, the lack of vaccine production inputs makes the vaccination against COVID-19 in Brazil take slow steps towards community immunization.

There is a worldly concern that the beginning of vaccination against COVID-19 may cause a decrease in the perception of the risk of infection [9]. The belief that immunization or recovering from a natural infection can promote full long-life protection is widely spread information among the population [10]. Besides, even after the strengthening of restrictions due to the explosive increase of cases and death toll after the holiday season, it is common to witness people-packed bars, streets, and even parties [11, 12]. More and more frequently, the news shows the lack of social distance between people, the absence of masks in public places, among other neglect of the population. Particularly in Brazil, the lack of a national plan to control the spread of SARS-CoV-2 and its variants has been jeopardizing the recovery of the economy and enhancing the health system’s fragility. Moreover, since the beginning of it, President Jair Bolsonaro has denied the importance of the epidemic, spreading fake news about the vaccine and encouraging the use of medications proven to be inefficient against COVID-19 [13, 14].

Brazil has started vaccination on priority groups, such as health professionals, older adults (80+), and indigenous people. But only about 2.7% of the Brazilian population has been achieved. Besides, Amazon state has been also prioritizing because of the emergence of new variants of the virus [15]. However, by the time vaccine is available for a broad population, acceptance and engagement are concerns. According to the Brazilian Immunization Society (SBIm) [16], there are three main reasons for the population to refuse vaccination: confidence, complacency, and convenience. In this context, we aimed to investigate, using an agent-based model (ABM), the vaccination’s impact under different scenarios of disease immunity, disease prevalence, individual behavior, and vaccination rate.

## 2 Methodology

### Mathematical Model

An extension of a previous agent-based model is proposed to assess the impact of vaccination against COVID-19 in Brazil [9]. The population is divided into eight epidemiological classes given the history of the disease and a campaigned vaccination in course, which are: susceptible; vaccinated; exposed; asymptomatic (and infectious); pre-symptomatic (and infectious); symptomatic with either mild or severe/critical illness, recovered; and dead. The model considers several types of heterogeneity, such as the contact matrix among age classes, the disease incubation and recovery period, the daily contact number among individuals, the percentage of individuals that show disease symptoms, the pre-existing herd immunity of each age class, prevalence of comorbidities in each age group, among others.

Every time-step, the number of daily contacts for each individual was sampled from a Poisson distribution based on their age group’s average number of contacts per day (see Table A.3). With probability *β*, a susceptible individual is infected after contacting an infected individual. *β* is dependent on the status of the infected individual (asymptomatic, pre-symptomatic, mild, or severe symptomatic). The incubation period was calculated from a Gamma distribution, based on an average estimate of 5.2 days. The time of incubation period in a pre-symptomatic stage is also calculated from a Gamma distribution. For symptomatic and asymptomatic individuals, the infectious period was also sampled from the Gamma distribution, with a mean of 3.2 days and five days, respectively. Depending on the age, symptomatic individuals can develop mild or severe conditions (see Table 1).

**Table 1:** Table of parameters.

| Description | 0–4 | 5–19 | 20–49 | 50–64 | 65–79 | 80+ | Source |
| --- | --- | --- | --- | --- | --- | --- | --- |
| Transmission probability per contact during presymptomatic stage | Depending on the level of herd immunity |  |  |  |  |  | Calibrated to R=1.04 [17] |
| Incubation period (days) | LogNormal(shape: 1.434, scale: 0.661) |  |  |  |  |  | [18] |
| Asymptomatic period (days) | Gamma(shape: 5, scale: 1) |  |  |  |  |  | Derived from [19, 20] |
| Presymptomatic period (days) | Gamma(shape: 1.058, scale: 2.174) |  |  |  |  |  | Derived from [21, 22] |
| Infectious period from onset of symptoms (days) | Gamma(shape: 2.768, scale: 1.1563) |  |  |  |  |  | Derived from [20] |
| Proportion of infections that are asymptomatic | 0.30 | 0.38 | 0.33 | 0.33 | 0.19 | 0.19 | [23–25] |
| Proportion of symptomatic cases that exhibit mild symptoms | 0.95 | 0.90 | 0.85 | 0.60 | 0.20 | 0.20 | [9] |
| Proportion of cases hospitalized with one or more comorbidities | 37.6% |  |  |  |  |  | [26, 27] |
| Non-ICU | 67% |  |  |  |  |  |  |
| ICU | 33% |  |  |  |  |  |  |
| Proportion of cases hospitalized without any comorbidities | 9% |  |  |  |  |  | [26, 27] |
| Non-ICU | 75% |  |  |  |  |  |  |
| ICU | 25% |  |  |  |  |  |  |
| Length of non-ICU stay (days) | Gamma(shape: 4.5, scale: 2.75) |  |  |  |  |  | Derived from [28, 29] |
| Length of ICU stay (days) | Gamma(shape: 4.5, scale: 2.75) + 2 |  |  |  |  |  | Derived from [28, 29] |

Mild symptomatic cases and severely ill cases isolate themselves within 24 hours once the symptoms started. We assumed (due to Brazil’s lack of data) that the number of contacts is reduced to a maximum of three, until recovery, on self-isolation. A proportion of the severe cases were hospitalized in a period of 3 to 9 days from the commencement of the symptoms [30], depending on their comorbidity status (see Table 1). The hospitalization period for ICU and ward was sampled from Gamma distribution, averaging 14.4 days and 12.4 days, respectively. Once hospitalized, the individual did not contribute to the spread of infection any longer. We assumed that long-life immunity is achieved after a successful recovery from an infection. The mortality probability is derived from [31], per age group, and applied to severe cases (see Table A.4).

We implemented the two-dose vaccination campaign already in the course in Brazil. The vaccines doses are distributed following the vaccination plan announced by the Health Minister of Brazil [32] and prioritizes the following groups (decreasing order): (i) health-care workers and the age group 80+; (ii) individuals that are between 60 to 79 years old; (iii) comorbid individuals in the age group 18 to 59 years old; (iv) general population. We assumed that vaccination takes place until the total coverage reaches 40% of the population, which is enough to keep vaccinating during the whole simulated epidemic peak (that takes around 200 days).

### Model’s parametrization

We used the effective reproduction number of 1.04, reported for the São Paulo State on January 10th [17], to calibrate the model and find the transmission probability per contact between a pre-symptomatic individual and a susceptible individual. The transmission probability of asymptomatic, mild, and severe symptomatic cases was considered to be 26%, 44%, and 89% in the pre-symptomatic stage. The population’s age distribution is based on the one reported in São Paulo State, encompassing ten age groups [33]. The contact distribution matrix is derived from Prem et al. [34], dividing the population into five age groups (see Table A.3). Besides, the presence of comorbidities is stratified by age (see Table A.1) [35].

Vaccine efficacy against symptoms is 50.38% for CoronaVac (from Sinovac) and 70.42% for ChAdOx1 nCoV-19 vaccine (from Oxford-AstraZeneca) [3, 4]. For CoronaVac, due to the lack of information about the single-dose efficacy, we supposed that the first dose provides half of the second dose efficacy. Besides, we also supposed that, following vaccination, the first and second doses’ efficacy is established after 14 and 7 days, respectively. For ChadOx1, the first dose efficacy is 64.1%, and each dose’s protection is established after 21 and 14 days after the vaccination. Since there is no statistical significance of protection against severe symptoms during phase 3 of both vaccines, we simulated two scenarios of protection against severe symptoms: no protection (0%) and complete protection (100%). Any other result must lay between them.

Regarding the protection against infection, we supposed three different scenarios: 0%, 50%, and 100% of the vaccine efficacy against symptoms. We supposed a vaccination rate of 0.3% of the population per day, which can be conservative given the vaccine production capacity reported both for CoronaVac and ChadOx1 in Brazil. We also simulated a scenario with a double vaccination rate, which may be achieved when combining both vaccines’ production capacity. Lastly, a low-risk perception scenario, where vaccinated individuals do not isolate themselves when they have mild symptoms, is analyzed.

The initial condition is given by one individual, in the pre-symptomatic stage, arriving at a population. We suppose a preexisting immunity in the population of 10, 20, and 30% [36]. Regarding the age distribution of population immunity, this was sampled following the reported age distribution of São Paulo State cases [31] (see Appendix, Table A.2). After, we relaxed the initial condition to assume a high prevalence of the disease. In summary, the baseline scenario encompasses isolation of symptomatic individuals and a vaccination rate of 0.3% of the population per day; in the double vaccination rate, the rate is increased to 0.6% per day; and in the low-risk perception, the rate is kept at 0.3%, and vaccinated individuals do not isolate themselves when having mild symptoms. The results shown are an average of 1000 independent Monte-Carlo realizations. The model was implemented in Julia language and is available at https://github.com/thomasvilches/abm_brasil.

## 3 Results

Let us define the “relative protection against infection” as the fraction of the vaccine efficacy against disease symptoms that is also applied against disease transmission. It means that, for instance, for the CoronaVac, as vaccine efficacy against symptoms is 50.38%, the scenario with 100% of relative protection means that protection against disease transmission is 50.38%; if the relative protection against infection is 50%, then protection falls to 25.19% against disease transmission; and if it is 0%, then there is no protection against disease transmission. The efficacy against severe symptoms is either 0% (no protection) or 100% (total protection), which means that none of the vaccinated individuals develops severe symptoms.

Firstly, we accessed the total number of infections, ward and ICU hospitalizations, and deaths resulting from each scenario of vaccine efficacy. Figures 1 and 2 display the results, supposing a previous herd immunity of 20% in the population. In the absence of a vaccine, it is generated almost 23,000 infections per 100,000 population. In a scenario of vaccination where vaccine provides the same level of protection against infection as against symptoms and complete protection against severe symptoms (scenario 100%-100% in Figures 1 and 2), the number of infections decreases to 22,140 for CoronaVac and 19,320 for ChadOx1. Notice that when the protection against infection is zero, the number of infections overcomes the no vaccination scenario for both vaccines (scenarios 0%-0% and 0%-100% in Figures 1 and 2). This happens because asymptomatic individuals do not isolate themselves, and vaccination protects against disease symptoms.

**Figure 1:**
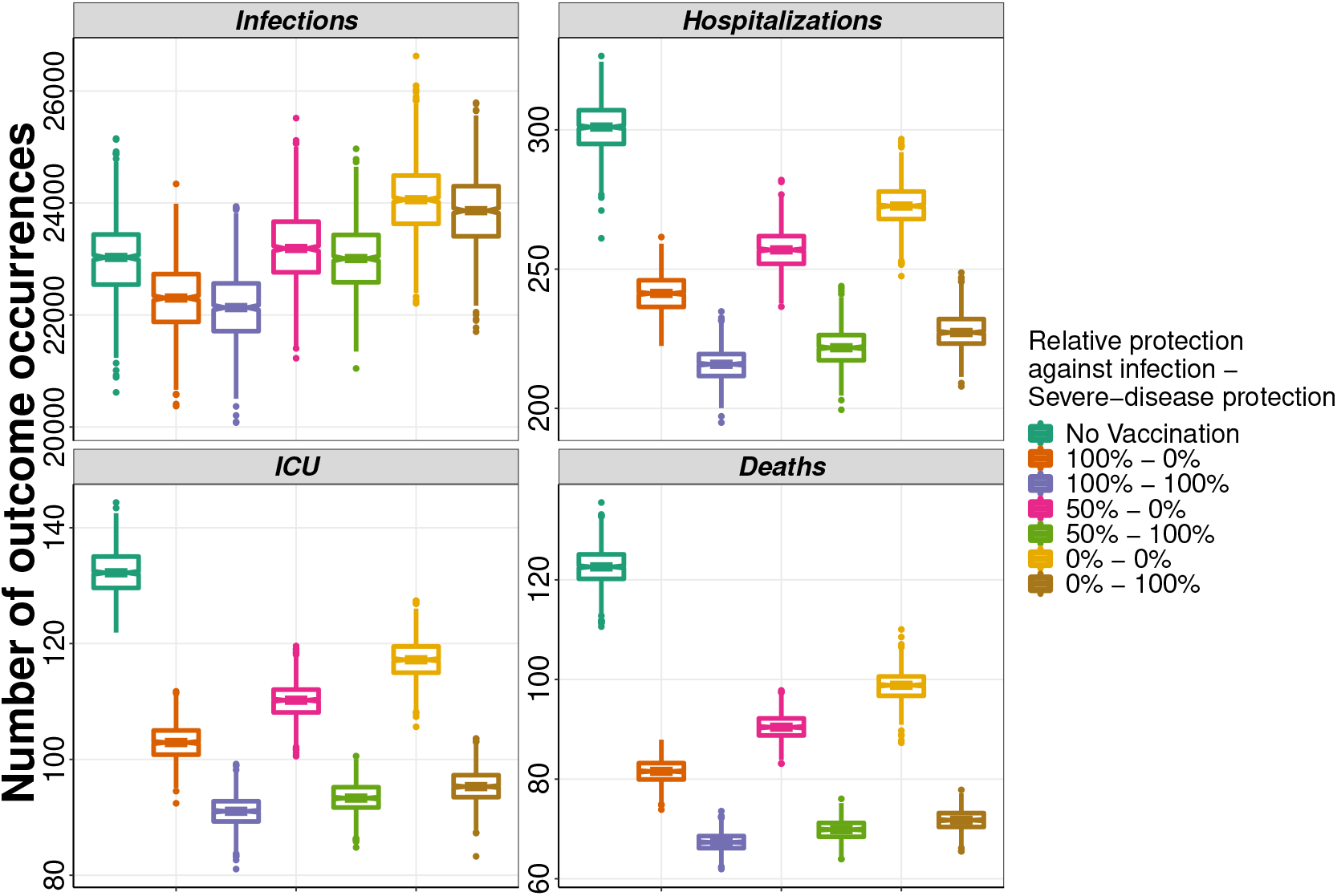
Outcomes for the baseline scenario and the CoronaVac vaccine. The second dose is given after 21 days.

**Figure 2:**
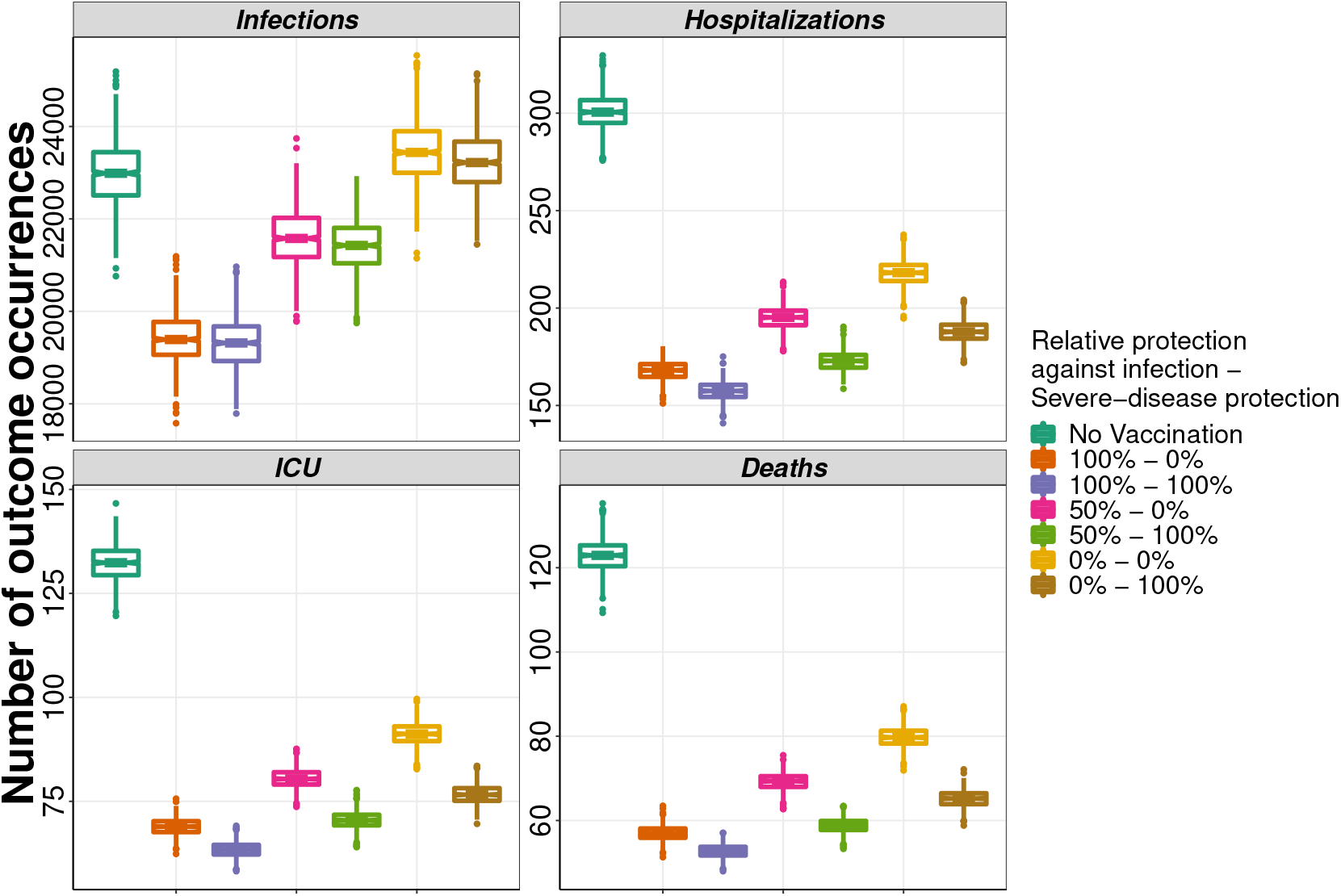
Outcomes for the baseline scenario and the AstraZeneca/Oxford vaccine. The second dose is given after 90 days.

It is important to highlight that even with a higher number of infections, vaccination may prevent many deaths and hospitalizations. Even in the case in which the vaccine does not provide any level of immunity against infection, the number of deaths decreases from 122 to 99 for CoronaVac and 80 for ChadOx1 when there is no protection against severe disease (scenario 0%-0% in Figures 1 and 2). The same patterns have been seen in hospitalizations (ward and ICU). This happens because although the vaccine does not prevent infections, it prevents symptoms and avoids possible severe outcomes of the disease.

Another way to compare the different scenarios is by measuring the percentage reduction in the average number of outcomes concerning the scenario in the absence of vaccination (Figures 3 and 4). In the baseline scenario, CoronaVac does not provide a significant reduction in the total number of infections in one epidemic peak, which takes around 200 days. However, it does prevent several of the disease outcomes. In the worst scenario of efficacy, in which the vaccine does not prevent either infection or severe symptoms (scenario 0%-0% in Figure 3), the vaccination may avoid 19.5% of deaths (95% CI: 12.79% −25.93%), 11.1% (95% CI: 2.92% −17.77%) of ICU admissions and 9.44% (95% CI: 2.18% −15.91%) of non-ICU hospitalizations. In the best case, when the vaccine prevents infection and symptoms equally, as well as it protects 100% against severe cases (scenario 100%-100% in Figure 3), a reduction of 45.2% (95% CI: 40.58% −49.66%), 28.19% (95% CI: 22.43% −33.99%), and 31.32% (95% CI: 25.25% −36.58%) can be achieved regarding the number of deaths, non-ICU hospitalizations, and ICU admissions.

**Figure 3:**
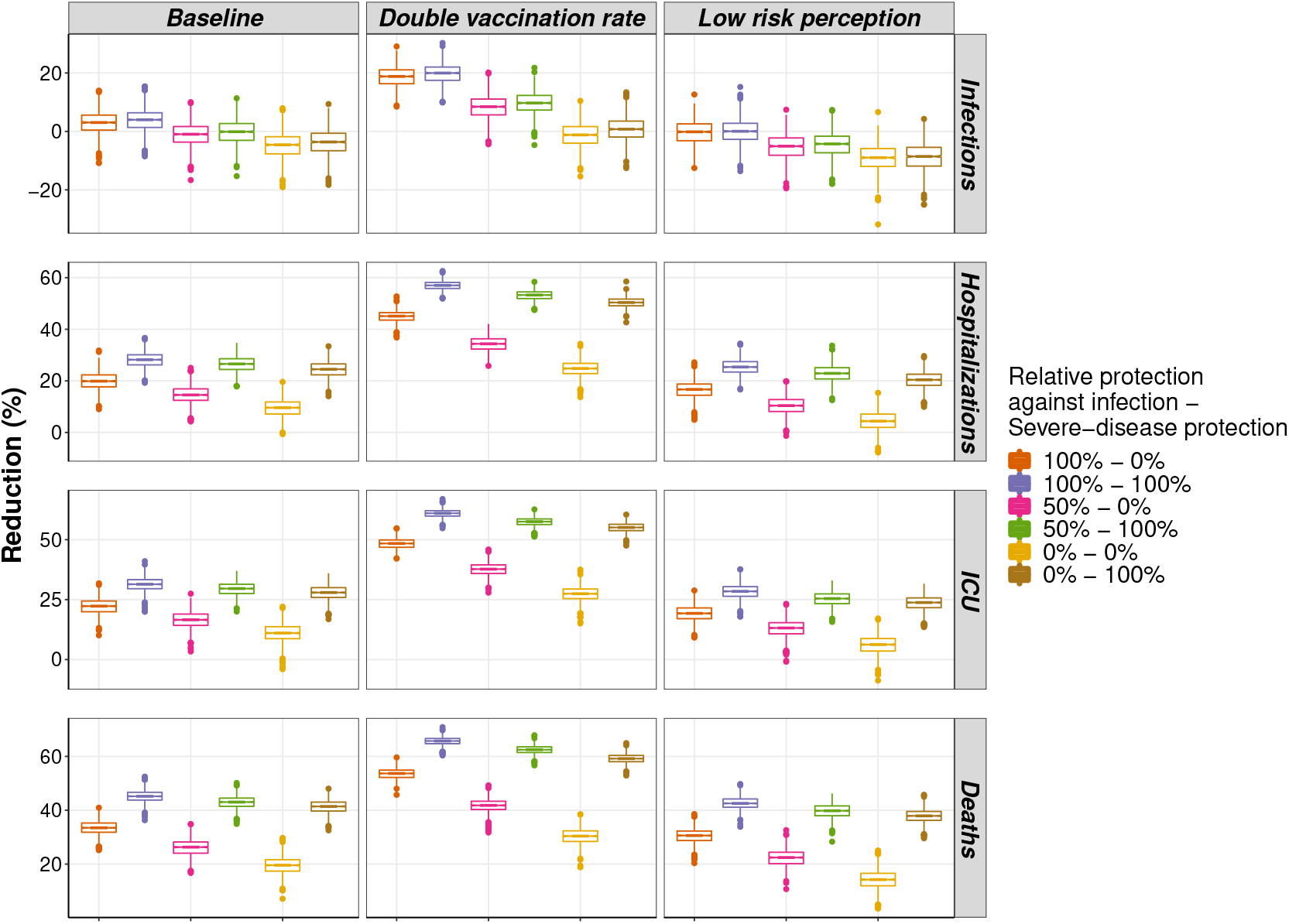
Percentage reduction in outcomes for the different scenarios and the CoronaVac vaccine. The second dose is given after 21 days.

**Figure 4:**
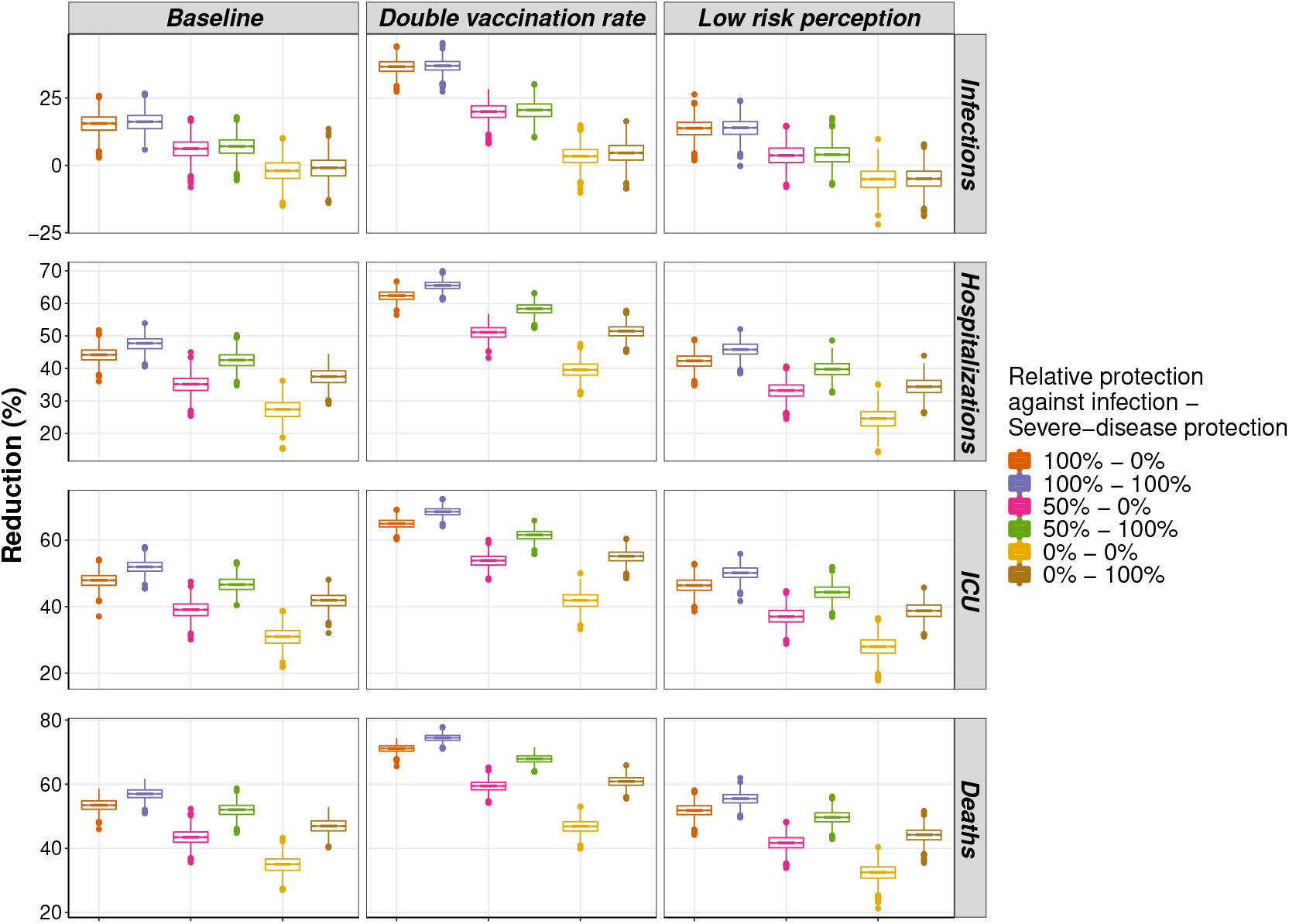
Percentage reduction in outcomes for the different scenarios and the As-traZeneca/Oxford vaccine. The second dose is given after 90 days.

Still in the baseline scenario, the vaccine ChadOx1 can perform even better (Figure 4), given its reported efficacy. In the worst efficacy assumption, in which the vaccine does not prevent against neither infections nor severe symptoms (scenario 0%-0% in Figure 4), the reductions in deaths, non-ICU hospitalizations and ICU admissions correspond to 34.98% (95% CI: 29.65% −40%), 27.31% (95% CI: 20.98% −33.75%) and 30.88% (95% CI: 25.15% −36.07%), respectively. When considering the best situation (scenario 100%-100% in Figure 4), ChadOx1 would prevent 56.98% (95% CI: 53.13% −60.41%) of deaths, 52.03% (95% CI: 48.24% −56.04%) of ICU admissions, and 47.59% (95% CI: 43.15% −51.57%) of non-ICU hospitalizations.

Doubling the vaccination rate with the CoronaVac, the worst scenario (scenario 0%-0% in Figure 3) presents a reduction of 30.25% (95% CI: 24.77% − 35.24%) in the number of deaths. The prevention of hospitalizations and ICU admissions more than doubled compared to the baseline scenario, reaching reduction values of 24.71% (95% CI: 18.66% - 30.15%) and 27.35% (95% CI: 20.85% - 33.09%), respectively. When it is considered that vaccine prevents against infection at the same level as it prevents against symptoms, and protects 100% against severe cases (scenario 100%-100% in Figure 3), the reduction of deaths would be 65.68% (95% CI: 62.64% - 68.56%). The ICU admissions would be reduced by 60.94% (95% CI: 57.39% - 64.08%), and non-ICU hospitalizations 57% (95% CI: 53.48% - 60.58%).

Considering a double vaccination rate for the ChadOx1 vaccine, the results are even better. The worst scenario (scenario 0%-0% in Figure 4) the number of deaths is reduced by 46.8% (95% CI: 42.37% - 50.81%), the non-ICU hospitalizations 39.59% (95% CI: 34.54% - 44.47%), and ICU admissions 41.87% (95% CI: 37.05% - 46.37%). In the best scenario (scenario 100%-100% in Figure 4), the reduction in the number of deaths exceeds 70%, whose value is 74.43% (95% CI: 72.1% - 76.55%). The prevention regarding the non-ICU hospitalizations is 65.54% (95% CI: 62.85% - 68.29%), while the prevention of ICU admissions is 68.57% (95% CI: 66.03% - 70.97%).

Nevertheless, when occurs a low-risk perception, the reduction values decrease. Considering the vaccine ChadOx1 in the best scenario (scenario 100%-100% in Figure 4), the reduction in the number of deaths, non-ICU hospitalizations, and ICU admissions decrease to 32.38% (95% CI: 26.72% - 37.47%), 24.58% (95% CI: 18.23% - 30.82%), and 27.95% (95% CI: 21.63% - 33.77%), respectively. While for CoronaVac (scenario 100%-100% in Figure 3), we reached the values of 43.59% (95% CI: 37.89% 47.11%), 25.4% (95% CI: 19.18% - 31.06%), and 28.35% (95% CI: 21.88% - 34.19%). In general, the decrease in the reduction ranges between 2% - 4%, revealing that if vaccinated people do not isolate themselves when mildly symptomatic, the vaccination result will be less efficient, which may correspond to thousands of lives. It shows the necessity of keeping other mitigation strategies while the process of vaccination occurs.

Finally, Figures 5 and 6 show the reduction in the outcomes when considering different initial infection prevalence scenarios. Instead of initializing the system with one pre-symptomatic individual, we variated the initial number of infections and assessed how it changed the vaccination’s impact (average reduction of disease outcomes).

**Figure 5:**
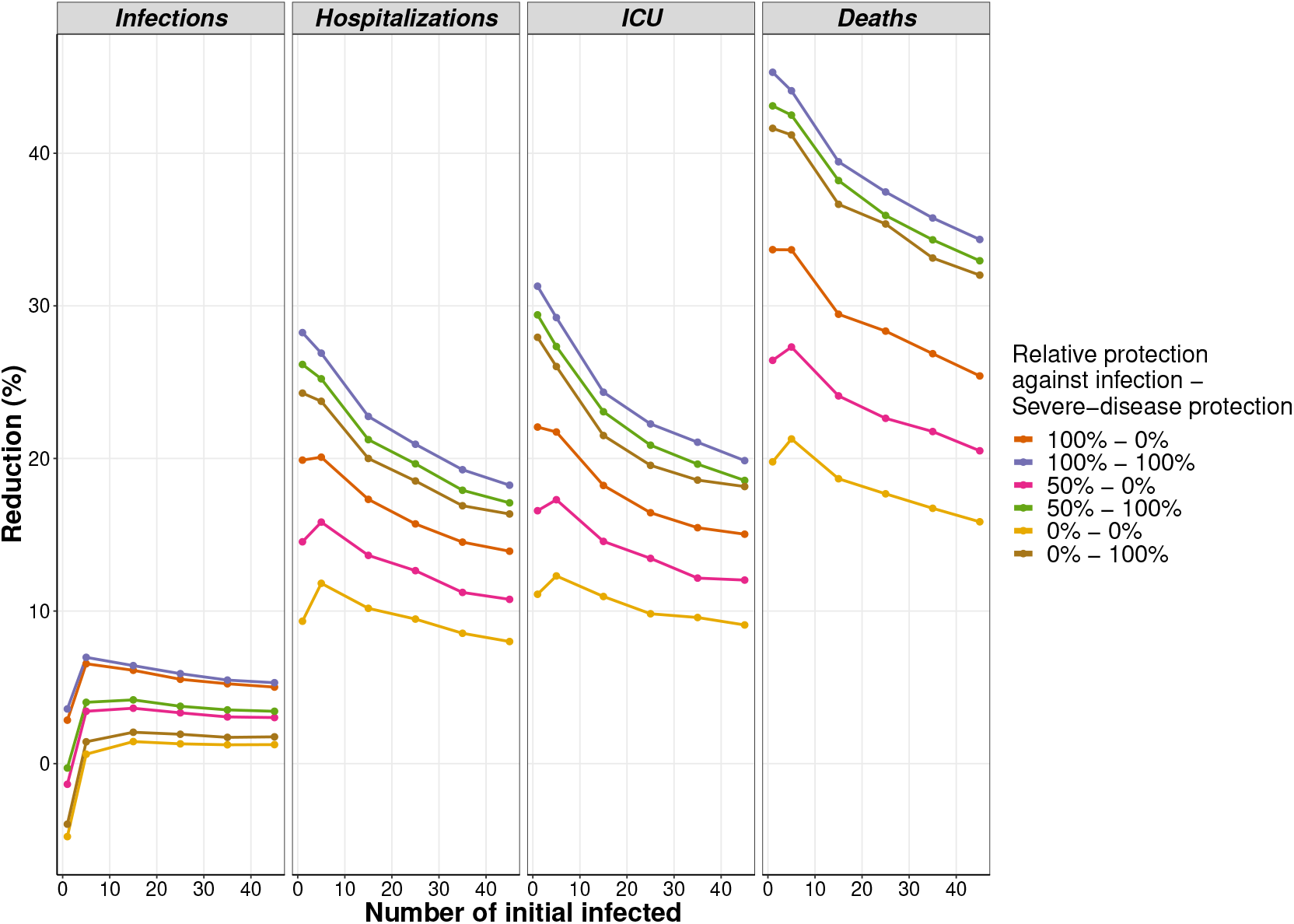
Average percentage reduction in outcomes in function of the initial number of infected individuals for the different scenarios and the CoronaVac vaccine. The second dose is given after 21 days.

**Figure 6:**
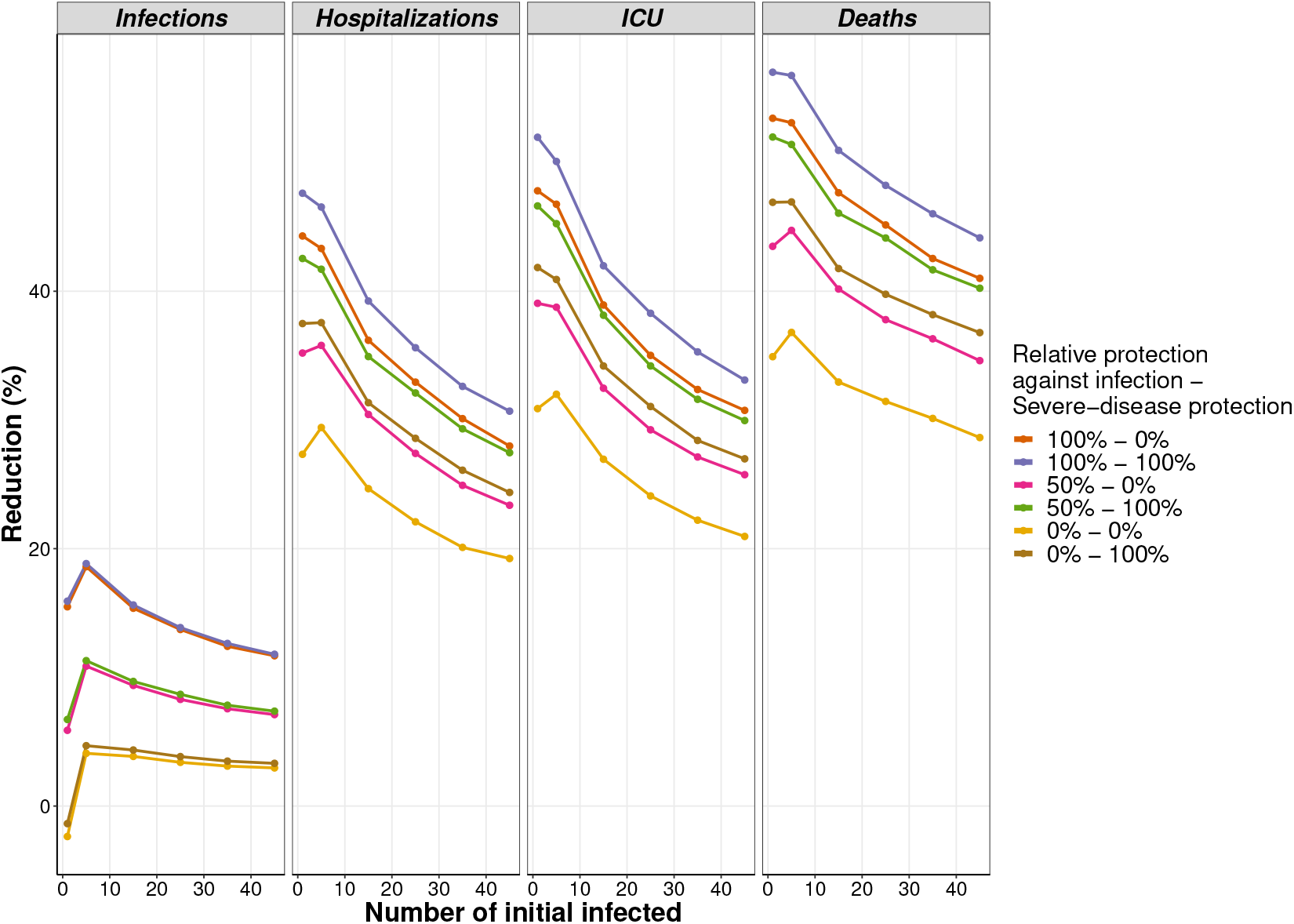
Average percentage reduction in outcomes in function of the initial number of infected individuals for the different scenarios and the AstraZeneca/Oxford vaccine. The second dose is given after 90 days.

Regarding the CoronaVac, considering the best-case scenario (scenario 100%-100% in Figure 5), concerning the number of infections, it was possible to observe an increase in the average reduction in outcomes when the initial number of infected individuals moves from one to five, from 3.58% (95% CI: −4.39% - 10.99%) to 6.96% (95% CI: 5.71% - 8.12%), followed by a decrease for higher number of initial infections, from 6.96% (95% CI: 5.71% 8.12%) to 5.30% (95% CI: 5.23% - 5.37%). In relation to non-ICU hospitalizations, the decrease in reduction was more drastic, going from 28.24% (95% CI: 22.27% - 34.12%) to 18.24% (95% CI: 17.88% - 18.58%). In turn, the ICU hospitalizations presented similar decrease in reduction comparing to the rearmost, going from 31.28% (95% CI: 25.78% - 36.50%) to 19.85% (95% CI: 19.36% - 20.30%). Again, the changes on the reduction in deaths are expressive, going from 45.30% (95% CI: 40.31% - 49.57%) to 34.35% (95% CI: 33.90% - 34.79%).

For the ChadOx1, increasing the initial prevalence of the disease, we obtain a significant decrease in the reduction of infections, non-ICU hospitalizations, ICU admissions, and deaths. In the best-case scenario (scenario 100%-100% in Figure 6), the reduction in the number of infections has a slight increase when changing the initial prevalence from one to five individuals, from 15.92% (95% CI: 9.19% - 22.55%) to 18.84% (95% CI: 17.66% 20.05%). For a initial prevalence of 45 infections, the reduction in infections jumps to 11.79% (95% CI: 11.71% - 11.88%). The non-ICU hospitalization reduction decreases from 47.62% (95% CI: 43.47% - 51.41%) to 30.68% (95% CI: 30.37% - 31%), while the ICU admission reduction was 51.97% (95% CI: 47.73% - 55.96%). The decline in the number of deaths is even more remarkable going from almost 60% (95% CI: 53.55% - 60.45%), for one initial infection, to 44.15% (95% CI: 43.76% - 44.54%), for 45 initial infections.

## 4 Discussion

We presented a stochastic agent-based model to simulate the SARS-CoV-2 transmission in a population and to discuss the scheme of vaccination already in the course in Brazil. The model is an extension of the one presented at [9], but it was parameterized with population characteristics of São Paulo State, Brazil. Besides, it explores the possibility of a behavior change caused by a decrease in risk perception. We analyzed the impact of the vaccination campaign supposing different vaccine efficacy scenarios, according to the two available vaccines in Brazil, the CoronaVac, from Sinovac-Butantan, and the ChadOx1, from AstraZeneca-Oxford-Fiocruz. Other key points that can jeopardize immunization, such as the population’s immunity, the disease prevalence, and the vaccination rate, were also accessed.

Important to note that Buss et al. [37] have scrutinized, through donated blood samples, the attack rate in Manaus and São Paulo, two of the most dramatic examples of the rapid spread of COVID-19 in Brazil [37]. The authors have found that in June (2020), one month after the epidemic peak in Manaus, 44% of the population had detectable immunoglobulin G (IgG) antibodies, with a 66% attack rate in June 2020 and 76% in October. The attack rate in São Paulo was 29% in October 2020. Specially Manaus was seen as an example of a city where herd immunity was almost achieved. Therefore new waves of the epidemic were not supposed to occur. Surprisingly, Manaus lives its worst epidemic moment now, with new variants of the virus emerging and spreading to other cities besides the health system’s collapse. This is a clear message that the only way to diminish the deaths, halt transmission, and recover the economy is through a global immunization program.

The impact of the vaccination campaign over disease control is not only related to the vaccine efficacy but also the velocity of vaccine production and distribution [38]. Competition about vaccine supplies is already in course, and low- and middle-income countries are more impacted by it. The baseline scenario was chosen to have a vaccination rate that corresponds to 0.3% of the population per day. Considering the Brazilian population, it would correspond to 640 thousand doses per day in the whole country. Moreover, we supposed vaccination campaign to be placed during the whole week, including weekends, which is already implemented in other countries, like the USA.

Given the reported number of vaccinated people per day during other immunization programs in Brazil, such as against poliomyelitis (which vaccinated 80 million children in three months) [8], it is fair to suppose that its production gives the limitation of vaccine distribution. In fact, the expected production rate of vaccines, when Butantan and Fiocruz reach their full capability, is around 1 to 3 million doses per day for each vaccine [5]. Therefore, our results are conservative in this aspect since it is also expected that Brazil can provide vaccine access to its neighborhoods countries in South America. The scenario with a double vaccination rate was able to decrease the disease outcomes highly. Even for CoronaVac, which has a lower reported efficacy compared to ChadOx1, in the situation that the vaccine does not prevents infections neither severe symptoms (scenario 0%-0% Figure 4), the reduction in ICU admissions and deaths was above 25%. This could take the health system off the borderline of collapsing. Today, most non-elective surgeries are canceled, prejudicing a lot of Brazilian patients that have their quality of life diminished by it. A fast vaccination would also stop the emergence and spread of new variants of the virus, which is already on course in Brazil, England, and Africa. Once fewer people get infected, the lower is the mutation probability of the virus.

When comparing the baseline scenario to the scenario in which the risk perception of vaccinated individuals is reduced (third column of Figures 3 and 4), we can see that 2% to 4% of the vaccination efficiency is lost, which can correspond to thousand of lives when looking at the number of deaths per 100 thousand population. This shows the importance of keeping mitigations strategies and an information campaign taking place together with vaccination. According to the Brazilian Immunization Society (SBIm) [16], the vaccination coverage by the PNI has steeply decreased since 2015, and in 2019 and 2020, none of the immunization programs has reached their goals. This is due to the lack of investments and good governance of the SUS services. As an example, the poor vaccination coverage has caused the once eradicated measles to reappear in Brazil. Another example is the vaccine for human papillomavirus (HPV). Santos et al. [39] show that despite 96.6% of the studied population took the vaccine, 32.3% did not take the second dose, which is considered a high dropout rate.

This study shows that the vaccines are keystones to preventing high attack rates and high tolls of hospitalizations and death, even when they provide limited protection against infection. The findings presented in our study are best interpreted by addressing the assumptions and limitations of this investigation. It was particularly challenging to find the data on the number of hospitalized individuals stratified by non-ICU and ICU types, as well as the steps addressed by the current national plan of immunization (PNI). Assessing the collective or individual behavior after the vaccination was not possible at the moment. We assumed that the PNI was able to vaccinate the vast majority of high-risk individuals, made up of 90% of healthcare workers and 70% of comorbid individuals below 65 years old, and 80% of older adults. It was also assumed that the population took both doses and that the daily number of contacts in the model was age-dependent but not activity-dependent. This means that we did not consider if daily contact was at the school, grocery stores, shopping malls, open areas, or any other places. Moreover, due to the lack of information about isolation in Brazil, we assumed that isolated people have a maximum of three contacts per day. The model also did not explicitly simulate other mitigation measures, such as mask use and social distance. Nonetheless, the model was calibrated using the current effective reproduction number. Therefore, it is expected to reflect the current mitigation measures in place.

This study presents the substantial gains of the vaccination program and its capacity for protecting vulnerable individuals. It is clear that once the public health actors apply adequate resources to ensure the proper mobility and action within the PNI framework, vaccination shall achieve the proposed goal of controlling outbreaks and lowering the disease burden. Fake news and behavioral change can diminish the vaccination rate and the population’s adherence to immunization. Therefore, a national and unique informative campaign addressed to the population is urgent.

## 5 Conclusion

Our findings support the immunization strategy proposed by the PNI to mitigate the disease burden and its societal impacts. It is also clear that all non-pharmaceutical efforts must continue to prevent the numbers from rising or remain at the same level. On the other hand, if the vaccination program triggers a behavior change in the population this can lead to the program’s lower efficiency as a whole. In this matter, we conclude that vaccines, along with non-pharmaceutical measures and a widely campaigned immunization, are key to the control of COVID-19 in Brazil.

## Data Availability

All the data is publicly available, including the source code of the model.

## Acknowledgments

We thank Professor Seyed M. Moghadas for the profitable discussion and comments, and for allowing us to use the computational resources from ABM-Lab, York University.

## A Supplementary information

The age distribution of the *in-silico* population is taking from the Brazilian Institute of Geography and Statistics [33], and it comprises seventeen age groups. Figure A.1 shows the age distribution for São Paulo State.

**Figure A.1:**
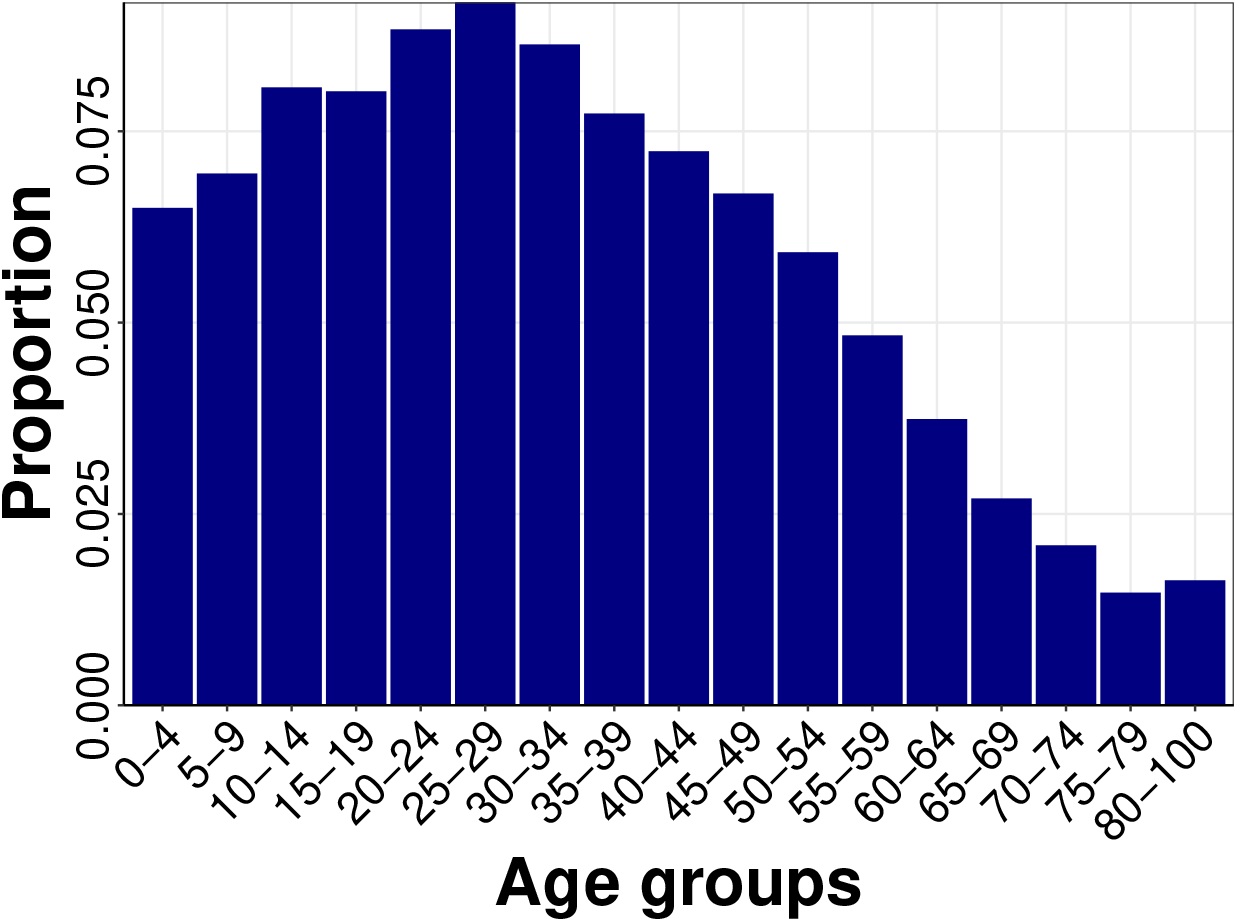
Population distribution per age group in São Paulo State [33].

The prevalence of comorbidities related to COVID-19 per age group is found at [35]. Table A.1 presents the percentage of the population that presents one or more comorbidities. We considered the sex ratio (female/male) as 0.5.

**Table A.1:** Prevalence of comorbidity in the population [35].

| Sex | Age groups |  |  |  |  |  |  |  |  |  |  |
| --- | --- | --- | --- | --- | --- | --- | --- | --- | --- | --- | --- |
|  | 0-4* | 5-17* | 18-24 | 25-29 | 30-34 | 35-39 | 40-44 | 45-49 | 50-54 | 55-59 | 60+ |
| Female | 5.0% | 10.0% | 12.2% | 14.5% | 23.3% | 27.7% | 37.5% | 46.6% | 52.1% | 62.2% | 75.0% |
| Male | 5.0% | 10.0% | 15.7% | 21.0% | 22.7% | 31.4% | 37.2% | 43.0% | 54.6% | 60.6% | 73.0% |

\* Value is taken from reference [40].

The preexisting herd immunity is distributed according to the reported distribution of cases per age class in São Paulo State, found at [31]. Table A.2 shows the probability of a pre-existing case to belong to a given age group.

**Table A.2:** Distribution of pre-existing herd immunity in the population per age class (%) [31].

| Age groups |  |  |  |  |  |  |  |  |
| --- | --- | --- | --- | --- | --- | --- | --- | --- |
| 0-9 | 10-19 | 20-29 | 30-39 | 40-49 | 50-59 | 60-69 | 70-79 | 80 |
| 2.6 | 4.5 | 9.1 | 14.7 | 20.3 | 23.6 | 17.5 | 5.2 | 2.5 |

The distribution of contacts between age groups, as well as the average number of daily contacts, is derived from [34]. Table A.3 shows the used distribution and the average number of daily contacts per age group.

**Table A.3:** Mixing patterns among individuals and the daily number of contacts derived from [34]. Daily numbers of contacts were sampled from Poisson distributions.

| Age groups |  | Proportion of contacts between age groups |  |  |  |  | Average number of daily contacts |
| --- | --- | --- | --- | --- | --- | --- | --- |
|  |  | 0-4 | 5-19 | 20-49 | 50-64 | 65+ |  |
| Individual's group | 0-4 | 0.285 | 0.176 | 0.405 | 0.098 | 0.035 | 11.80 |
|  | 5-19 | 0.035 | 0.644 | 0.256 | 0.050 | 0.015 | 21.32 |
|  | 20-49 | 0.043 | 0.163 | 0.652 | 0.119 | 0.022 | 15.80 |
|  | 50-64 | 0.060 | 0.170 | 0.529 | 0.203 | 0.038 | 12.71 |
|  | 65+ | 0.064 | 0.266 | 0.388 | 0.161 | 0.121 | 6.33 |

The probability of death in a symptomatic case was taken from [31], and it is shown in Table A.4 for each age group. Since we are considering that only severe cases can die, a formula is applied to calculate the death probability in severe cases (*ρ*_*sev*_):

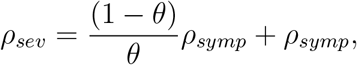

in which the *ρ*_*symp*_ is the death probability of a symptomatic case and *θ* is the probability that a symptomatic case is severe.

**Table A.4:** Percentage of symptomatic fatal cases per age group

| Age groups | 0-9 | 10-19 | 20-29 | 30-39 | 40-49 | 50-59 | 60-69 | 70-79 | 80-89 | 90+ |
| --- | --- | --- | --- | --- | --- | --- | --- | --- | --- | --- |
| Percentage of fatal cases | 0.1% | 0.1% | 0.1% | 0.4% | 1.0% | 2.7% | 8.1% | 17.7% | 30.8% | 37.9% |

## B Percentage reduction on disease outcomes varying the preexisting immunity

We present the obtained results for CoronaVac and AstraZeneca/Oxford vaccines for several scenarios: (I) baseline, doubling the vaccination rate, and decreasing the risk perception.

Figures B.1 and B.2 show the results considering a pre-existing immunity of 10%, for CoronaVac and AstraZeneca/Oxford vaccines, respectively. And Figures B.3 and B.4, a pre-existing immunity of 30%.

**Figure B.1:**
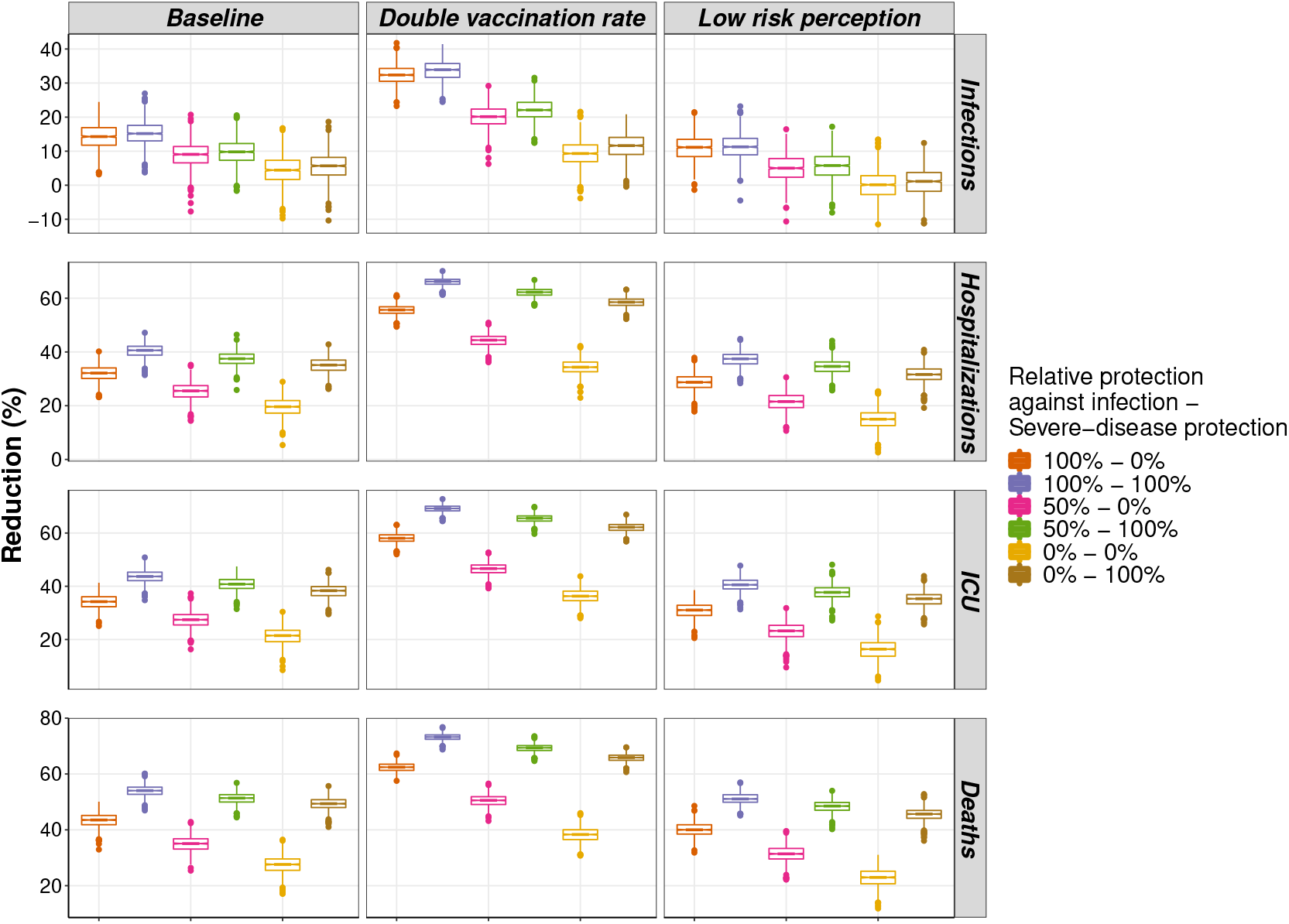
Percentage reduction in outcomes for the different scenarios and the CoronaVac vaccine, with pre-existing immunity in the population of 10%. The second dose is given after 21 days.

**Figure B.2:**
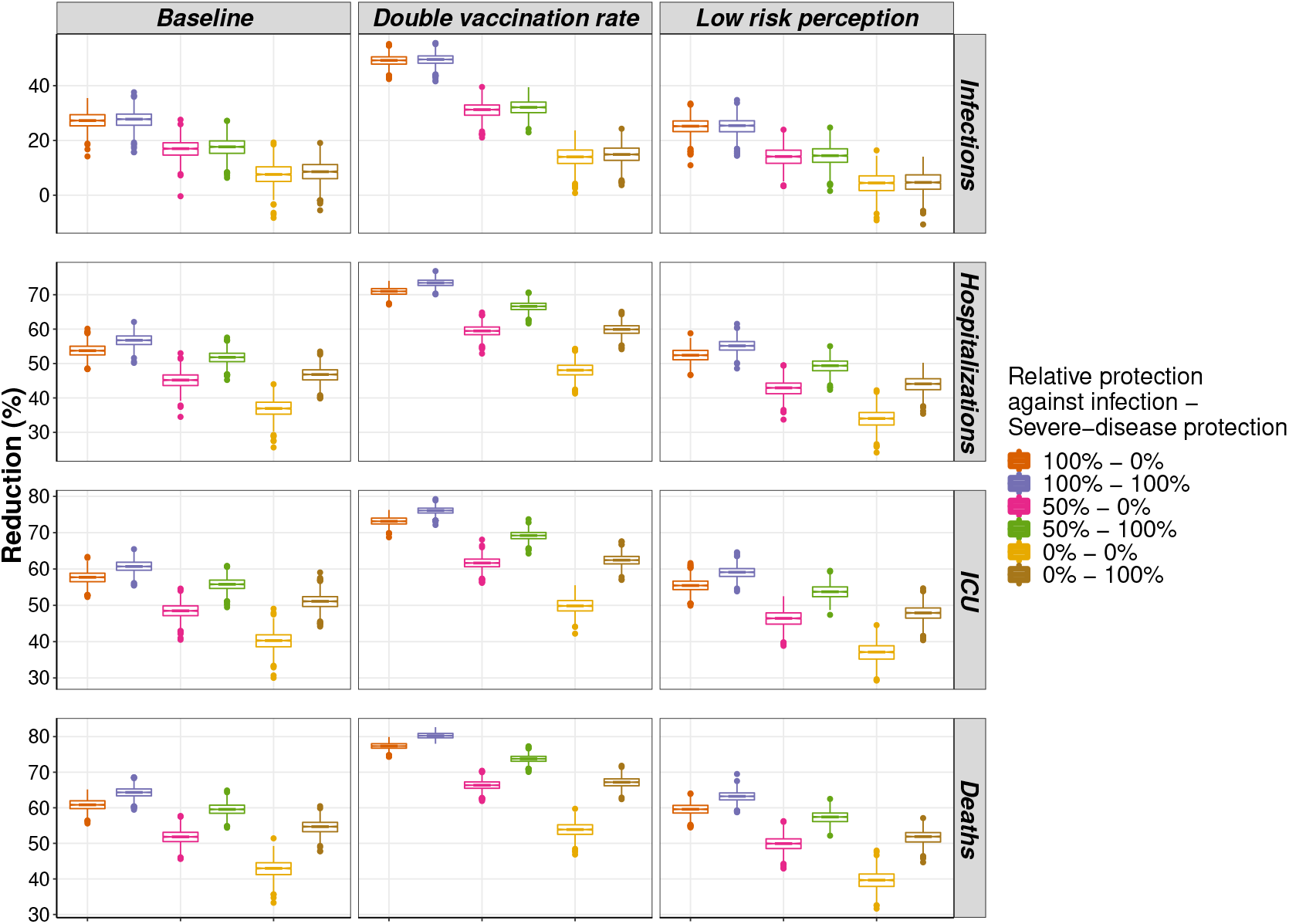
Percentage reduction in disease outcomes for the different scenarios and the AstraZeneca/Oxford vaccine, with pre-existing immunity in the population of 10%. The second dose is given after 90 days.

Tables B.1 to B.6 show the percentage of reduction in the number of infections, non-ICU hospitalizations, ICU admissions, and deaths, considering different levels of pre-existing immunity. The listed values summarize the results shown in Figures B.1 to B.4 and also the results in the main text.

**Figure B.3:**
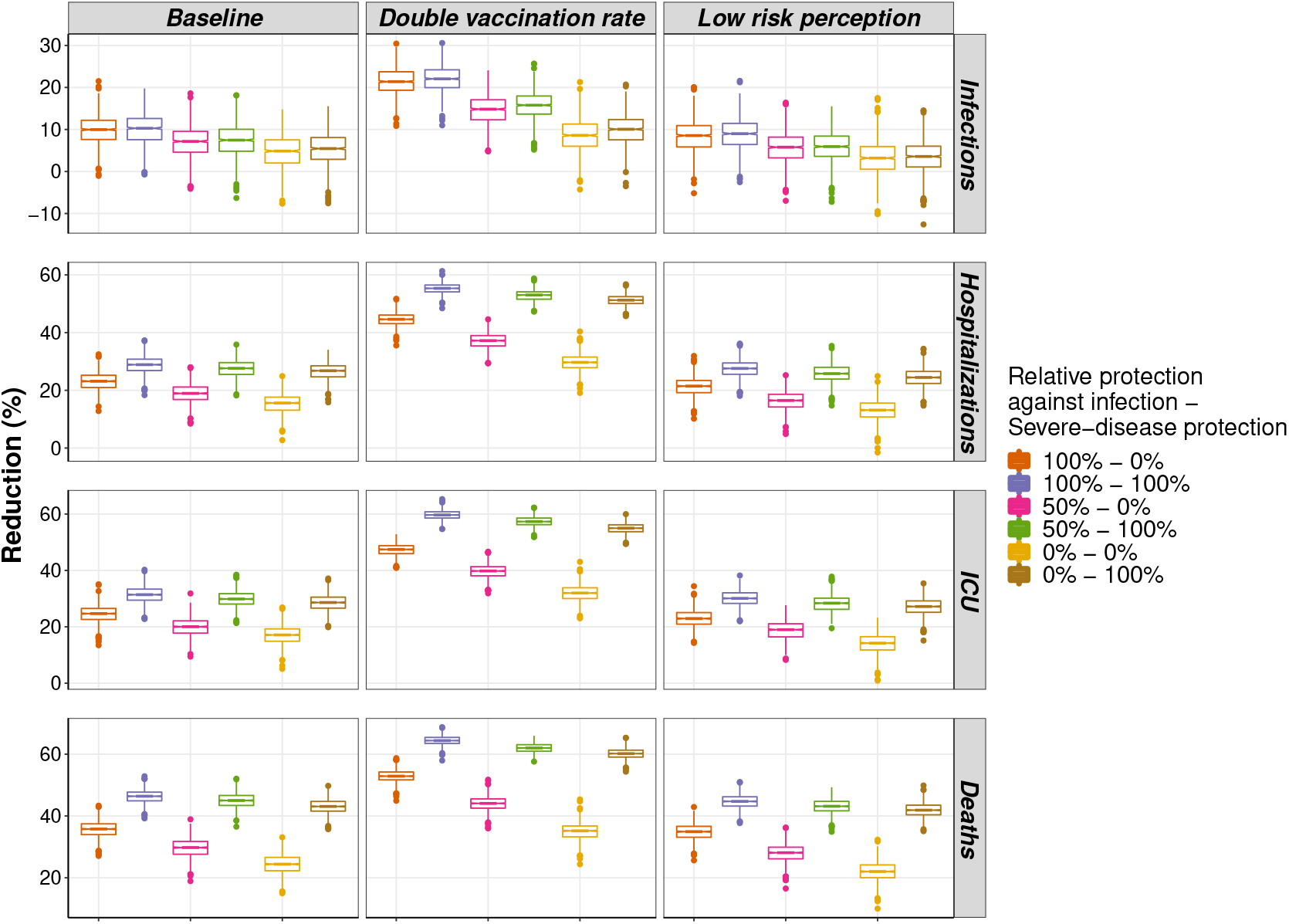
Percentage reduction in disease outcomes for the different scenarios and the CoronaVac vaccine, with pre-existing immunity in the population of 30%. The second dose is given after 21 days.

**Figure B.4:**
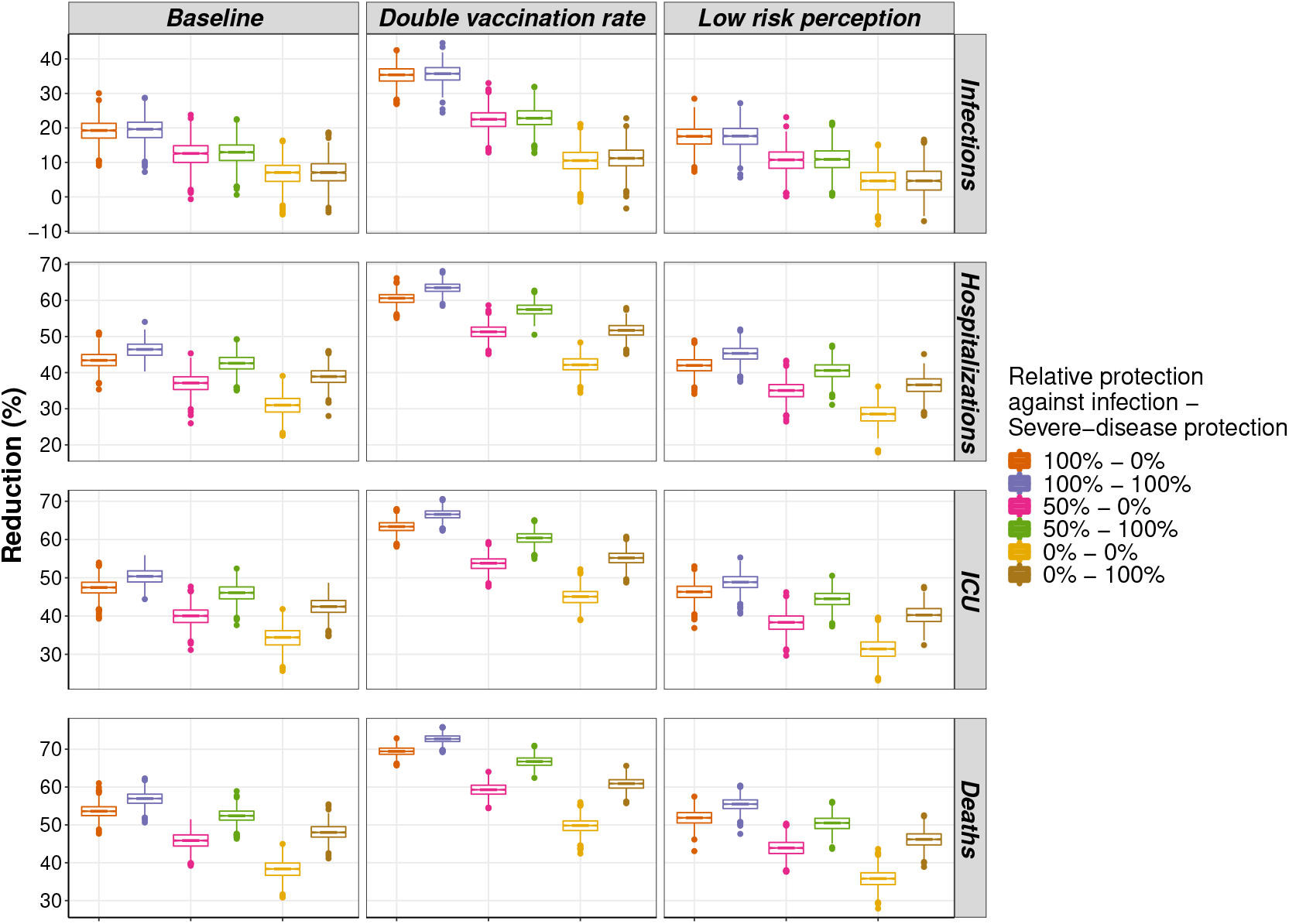
Percentage reduction in disease outcomes for the different scenarios and the AstraZeneca/Oxford vaccine, with pre-existing immunity in the population of 30%. The second dose is given after 90 days.

**Table B.1:**
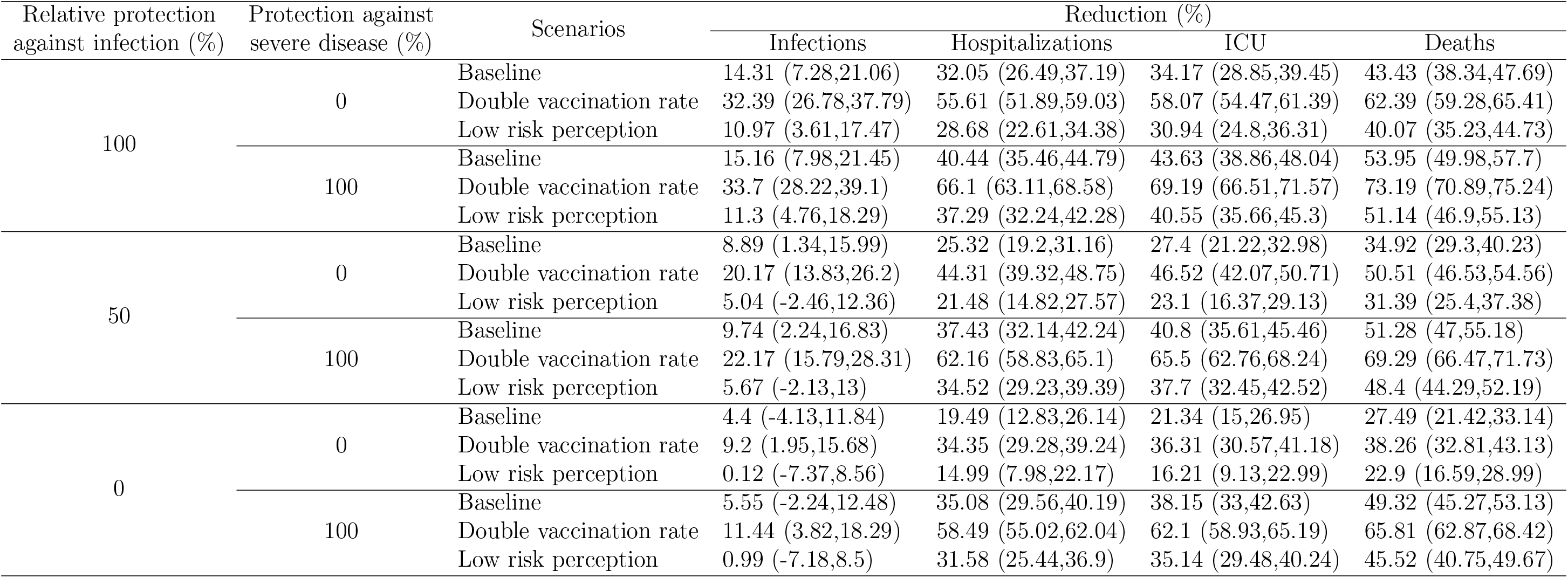
Percentage reduction in disease outcomes under different protection scenarios of the CoronaVac vaccine and pre-existing immunity in the population of 10%.

**Table B.2:**
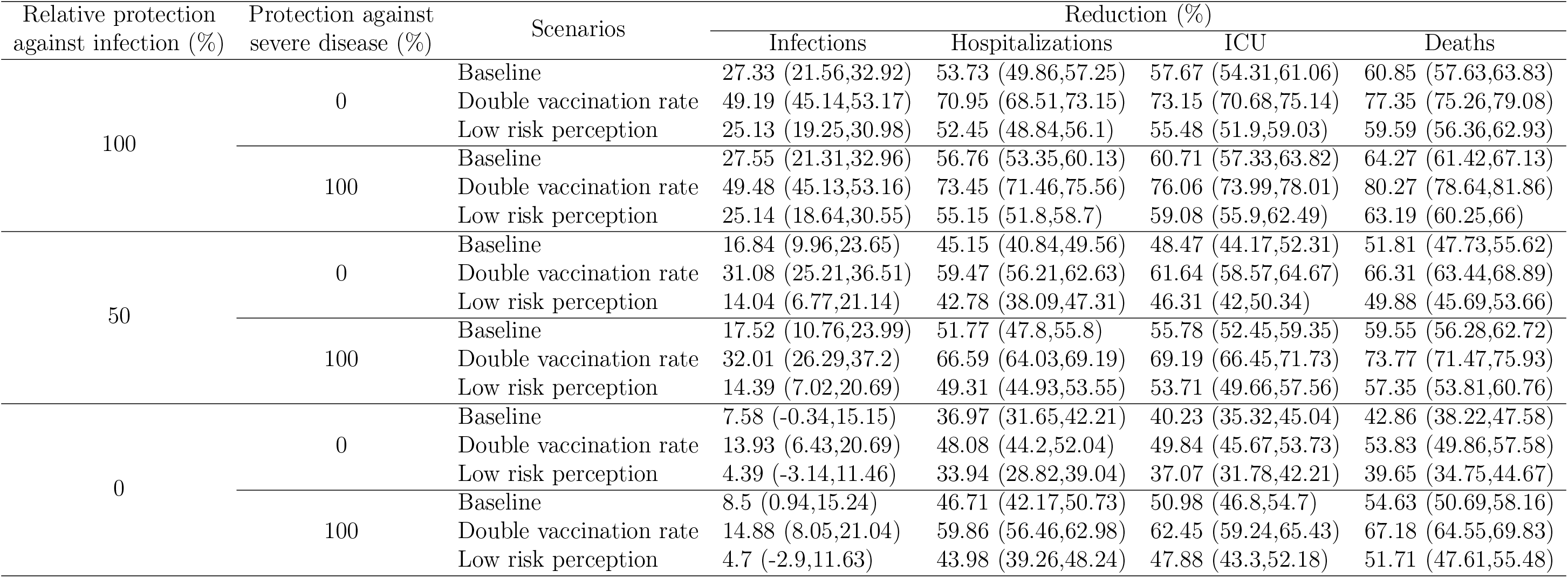
Percentage reduction in disease outcomes under different protection scenarios of the AstraZeneca/Oxford vaccine and pre-existing immunity in the population of 10%.

**Table B.3:**
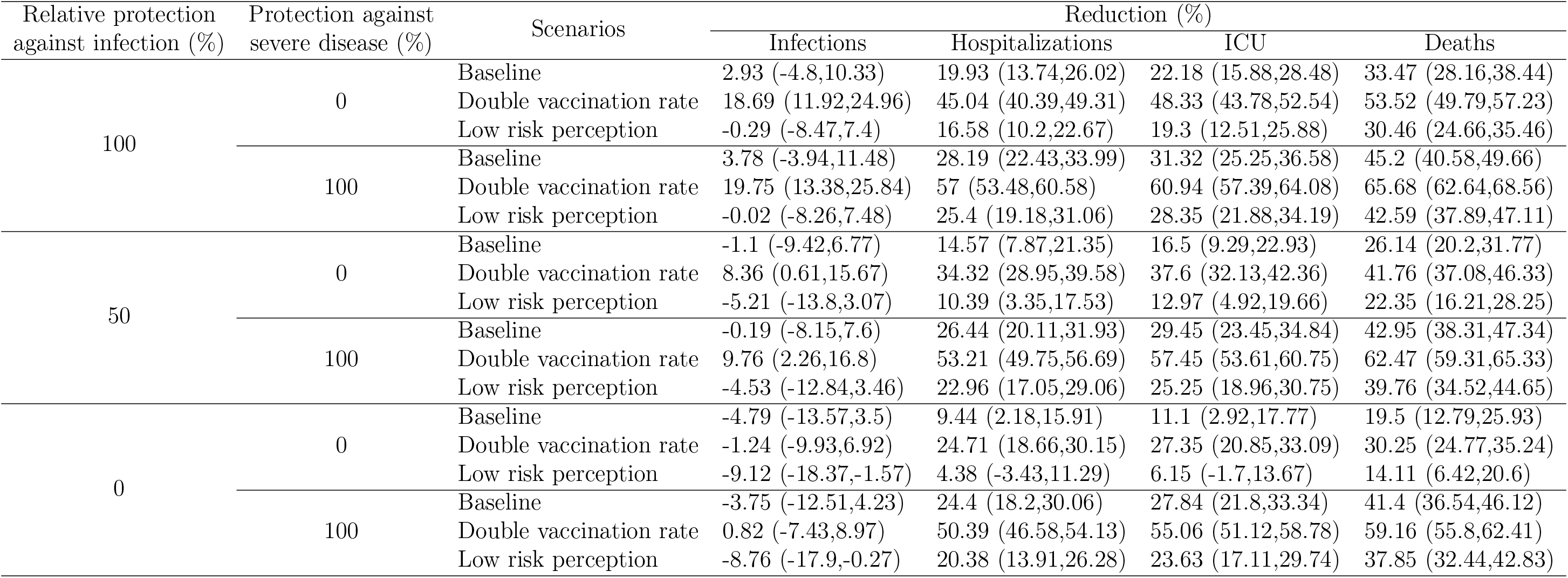
Percentage reduction in disease outcomes under different protection scenarios of the CoronaVac vaccine and pre-existing immunity in the population of 20%.

**Table B.4:**
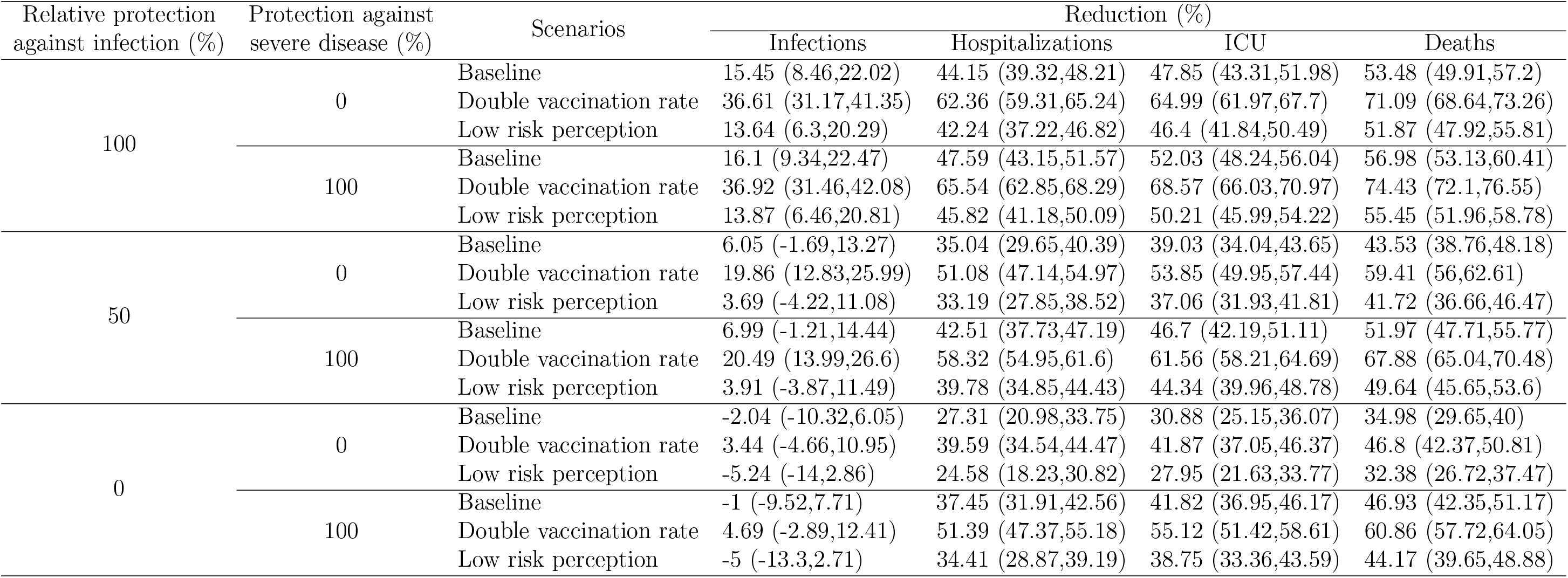
Percentage reduction in disease outcomes under different protection scenarios of the AstraZeneca/Oxford vaccine and pre-existing immunity in the population of 20%.

**Table B.5:**
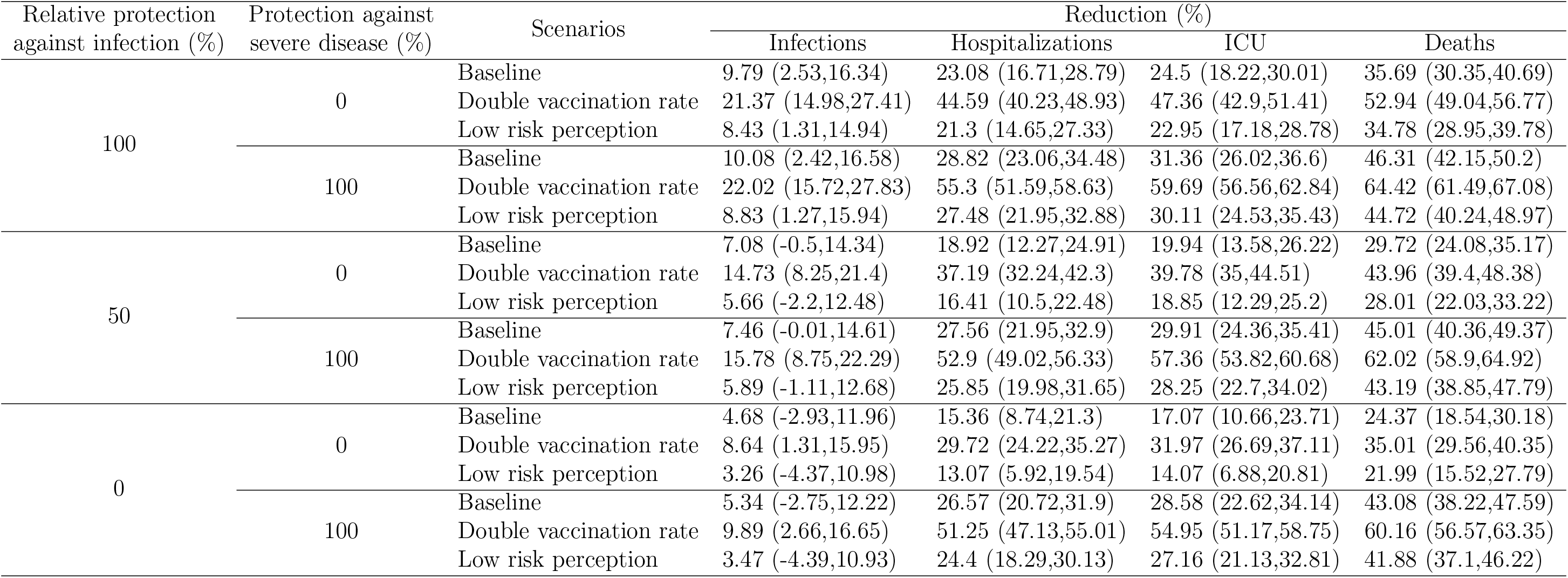
Percentage reduction in disease outcomes under different protection scenarios of the CoronaVac vaccine and pre-existing immunity in the population of 30%.

**Table B.6:**
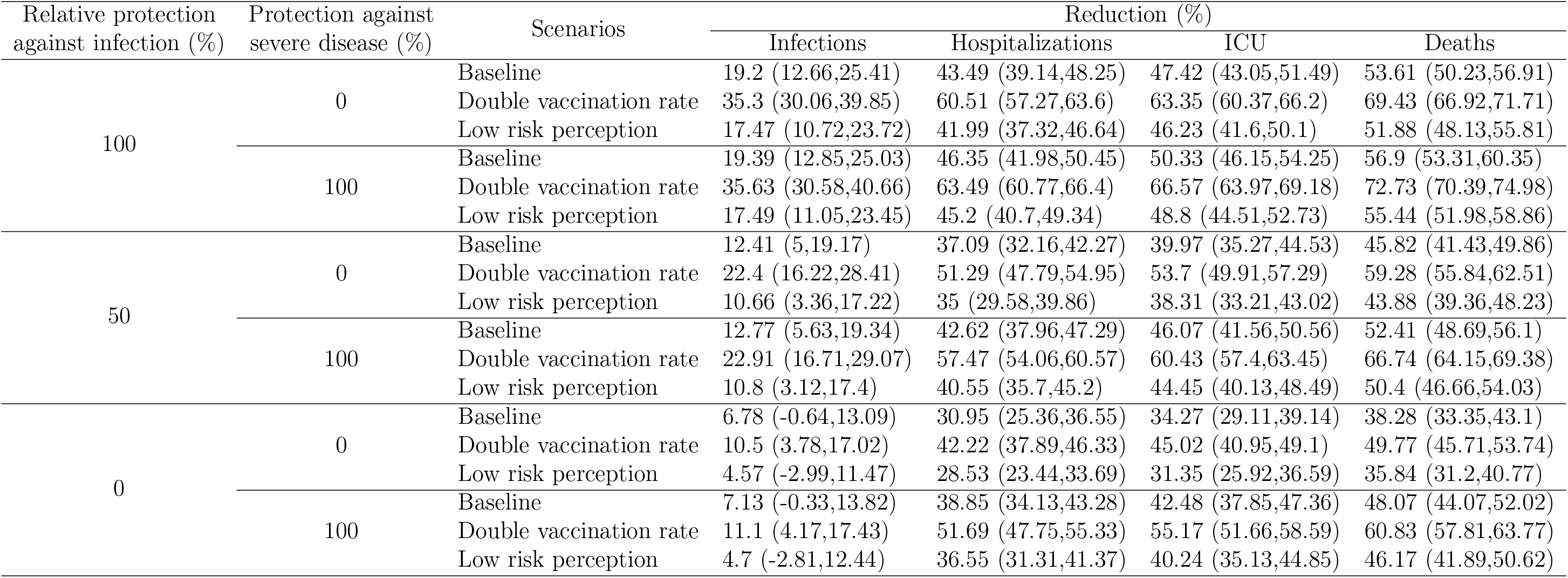
Percentage reduction in disease outcomes under different protection scenarios of the AstraZeneca/Oxford vaccine and pre-existing immunity in the population of 30%.

## Footnotes

1 Communities that used to be a slave settlement. Nowadays, they are protected areas in which the descendant of those slaves live.

